# Wearable Evidence Linking Dyskinesia Burden to Sleep Quality in Parkinson’s Disease

**DOI:** 10.64898/2026.05.30.26354503

**Authors:** Viktoria Azoidou, Essa Bhadra, Ellen Camboe, Kamalesh C. Dey, Alexandra Zirra, Kira Rowsell, Corrine Quah, Caroline Budu, Thomas Boyle, David Gallagher, Jonathan P. Bestwick, Laura Pérez-Carbonell, Alastair J Noyce, Cristina Simonet

## Abstract

**Background:** Sleep disturbances affect up to 60-80% of people with Parkinson’s disease (PD) and are associated with worse clinical outcomes and reduced quality of life. Dyskinesia is a common motor complication of dopaminergic therapy, but its relationship with sleep quality remains incompletely defined.

**Methods:** Forty-seven people with PD (median age 68 years; 44.7% female; median disease duration 5 years; 38.3% from non-White ethnic background) were assessed for sleep quality on Pittsburgh Sleep Quality Index (PSQI). Dyskinesia was assessed using Movement Disorder Society-Unified Parkinson’s Disease Rating Scale (MDS-UPDRS) Part IV items 4.1 and 4.2, and 7-day wearable monitoring with the Parkinson’s KinetiGraph (PKG) to derive median dyskinesia score (DK_50) and fluctuation dyskinesia score (FDS). All analyses were conducted using multivariate regression. Associations with sleep quality were adjusted for age, sex, and disease severity (MDS-UPDRS Part III) in Model A; additionally for levodopa equivalent daily dose (LEDD) in Model B; and further for disease duration in Model C.

**Results:** In Model A, all four dyskinesia measures were significantly associated with sleep quality. After adjusting for LEDD in Model B, only DK_50 remained a significant predictor of worse sleep (B=0.18, 95CI: 0.003-0.357, P=0.047). With additional adjustment for disease duration in Model C, the association for DK_50 was attenuated (B=0.18, 95%CI: -0.001 to 0.356, P=0.051).

**Conclusions:** Wearable-derived continuous dyskinesia burden was independently associated with worse sleep quality, whereas clinician-rated dyskinesia was not, highlighting the added clinical value of objective motor monitoring in PD. Disease duration may partly confound this relationship. Larger prospective studies are warranted.

## Introduction

Sleep disturbance is among the most prevalent non-motor features of Parkinson’s disease (PD), affecting up to 80% of patients within five years of diagnosis^1^. A 2023 meta-analysis reported pooled prevalences of 46% for REM sleep behaviour disorder and 35% for excessive daytime sleepiness in patients with PD^2^, and a separate pooled estimate placed poor sleep quality at 58% across 9,382 patients^3^. Sleep disturbances in PD arise from neurodegeneration of sleep-wake regulatory circuits, dopaminergic medication effects, and nocturnal motor symptoms, and independently predict worse health-related quality of life^4,5^.

Levodopa-induced dyskinesia (LID) is a motor complication of treated PD. In the COPPADIS cohort of 672 patients, LID increased from 18.9% at baseline to 42.6% at 5 years^6^, and population-based data show that around 30% of levodopa-treated patients develop dyskinesia at a median of 4 years from levodopa initiation^7^. Dyskinesia development is increasingly understood as a function of disease duration rather than cumulative levodopa exposure^8,9^, reflecting maladaptive plasticity within corticostriatal circuits, specifically, loss of bidirectional synaptic plasticity and impaired depotentiation^10,11^.

Sleep and dyskinesia have been linked in both clinical and neurophysiological data. In a previous cross-sectional study, poor nighttime sleep was independently associated with dyskinesia in 425 PD patients^12^, and prospectively, a high PSQI score predicted incident dyskinesia at 36 months^13^. Impaired sleep-dependent synaptic downscaling, reflected in a blunted overnight slow-wave activity decline, has been proposed as a shared mechanism^14,15^.

However, all prior studies linking sleep quality to dyskinesia have relied on intermittent, clinician-rated assessments, which are known to underestimate motor burden in PD^16-18^. Brief clinic encounters capture motor behaviour only during the period of direct observation, and are particularly insensitive to activity occurring in the late-afternoon, evening, and nocturnal periods most relevant to sleep architecture. The Parkinson’s KinetiGraph (PKG) is a validated wrist-worn accelerometer that samples motor activity every two minutes over seven to ten continuous days. Its dyskinesia output has been validated against the Abnormal Involuntary Movement Scale to within the inter-rater limits of agreement of three movement disorder neurologists^18^ and identifies clinically unrecognised early morning akinesia^17^.

No prior study has compared continuous wearable-derived and intermittent clinician-rated dyskinesia as correlates of global subjective sleep quality within the same cohort. We therefore examined whether dyskinesia burden, assessed by both approaches, was associated with subjective sleep quality in people with PD.

## Methods

This cross-sectional study included 47 patients with idiopathic PD recruited from specialist outpatient clinics at Barts Health NHS Trust and Homerton Healthcare NHS Foundation Trust. PD was diagnosed according to Movement Disorder Society clinical diagnostic criteria^19^. All participants provided written informed consent, and the study was approved by the London-Dulwich Research Ethics Committee (23/PR/1526).

Subjective sleep quality was assessed using the PSQI, a validated measure of global sleep quality over the preceding month^20^. The PSQI total score was prespecified as the primary outcome. Motor complications were assessed using Part IV of the Movement Disorder Society-Unified Parkinson’s Disease Rating Scale (MDS-UPDRS), with dyskinesia quantified using item 4.1 (time spent with dyskinesias) and item 4.2 (functional impact)^21^. Objective motor data were obtained from the PKG, worn continuously for 7 days. The median dyskinesia score (DK_50) and Fluctuation Dyskinesia Score (FDS) were the wearable-derived measures of dyskinesia burden^18^.

Cognitive function was measured using the Montreal Cognitive Assessment (MoCA)^22^. Demographic and clinical variables also recorded were: age, sex, self-reported race/ethnicity^23^, disease duration from formal diagnosis (years), Hoehn & Yahr stage^24^, levodopa equivalent daily dose (LEDD), and MDS-UPDRS Parts I-III scores ((non-motor experiences of daily living, motor experiences of daily living, and motor examination, respectively)^21^.

Analyses were performed in IBM SPSS Statistics version 29. Continuous variables are presented as median (IQR), and categorical variables as frequencies and percentages. Because dyskinesia measures were non-normally distributed, associations between PSQI total and each dyskinesia variable were examined individually using Spearman’s rank correlations. Partial Spearman correlations were computed in three pre-specified hierarchical adjustment models: Model A adjusted for age, sex, and MDS-UPDRS Part III (motor examination), selected *a priori* as potential confounders. Model B added LEDD^25^ and Model C added disease duration. Models were ordered by temporal precedence (demographics, current motor severity, cumulative pharmacological exposure, and disease duration as a progression proxy). Multivariable linear regression was performed in parallel using the same Models A, B, and C, with PSQI total as the dependent variable. All tests were two-sided (α=0.05). No correction for multiple comparisons was applied given the exploratory design.

## Results

Forty-seven patients with PD were included (TABLE 1). Median age was 68 years (IQR 62-76); female sex, 44.7%; median disease duration, 5 (2-8) years; median MDS-UPDRS Part III, 45.0 (36.5-50.5), and median LEDD, 400 (300-500) mg/day. Dyskinesia severity was mild to moderate (median MDS-UPDRS Part IV items 4.1 and 4.2 score 0 [0-2]; PKG DK_50 1.40 [0.55-2.50]; FDS 8.10 [6.35-10.20]). Median PSQI was 10.0 (8.0-14.5), well above the validated cut-off of five^20^.

**TABLE 1.**
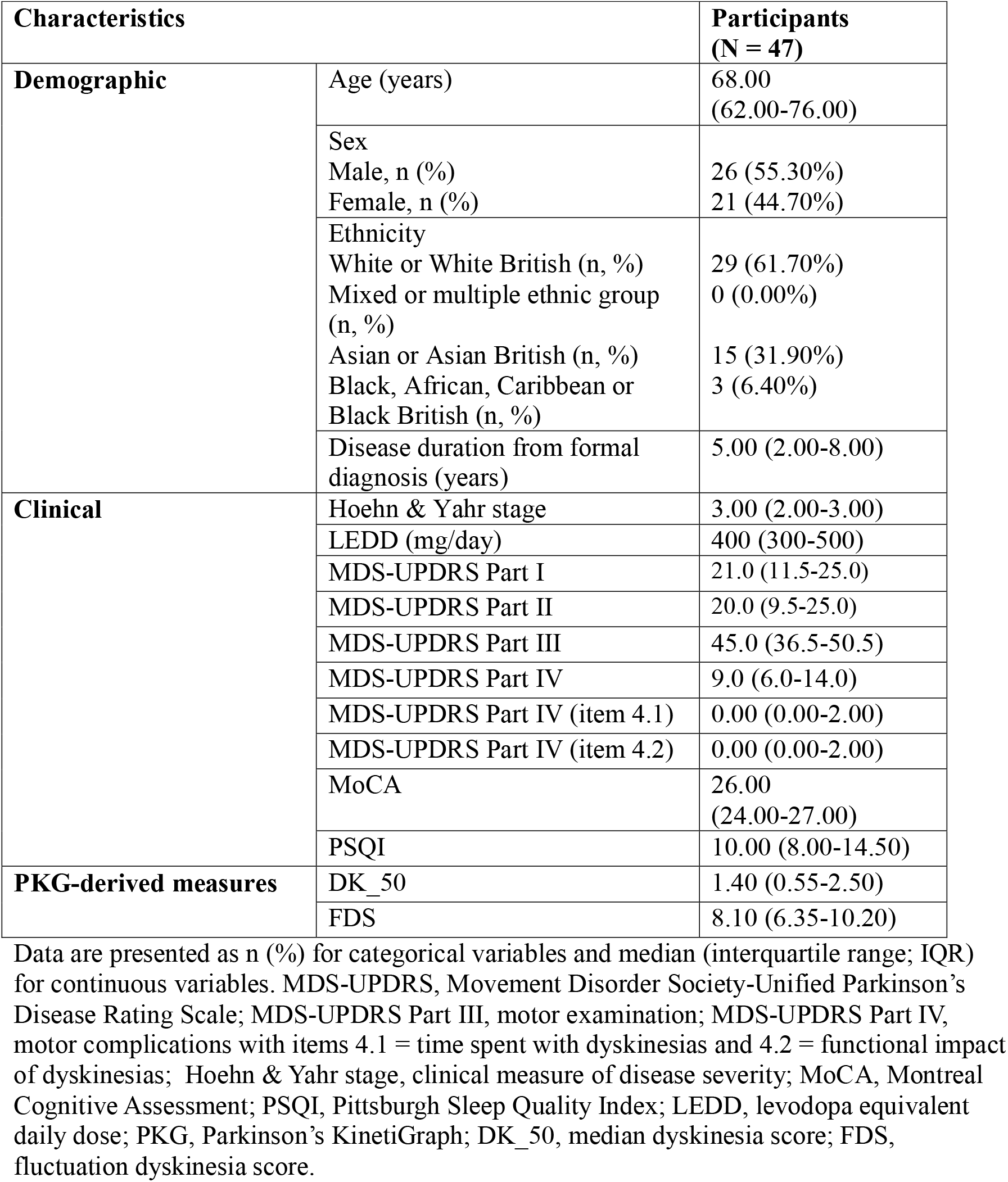
Demographic, clinical, and questionnaire characteristics.

In unadjusted analyses, worse subjective sleep quality (higher PSQI) was associated with greater dyskinesia burden across clinical and wearable-derived measures alike (SUPPLEMENTARY TABLE 1). PSQI correlated with MDS-UPDRS Part IV item 4.1 (ρ=0.386, P=0.007), item 4.2 (ρ=0.392, P=0.006), and DK_50 (ρ=0.487, P<0.001). The association with FDS did not reach significance (ρ=0.265, P=0.072).

After adjusting for age, sex, and motor examination (MDS-UPDRS Part III; Model A), the association between DK_50 and PSQI strengthened (ρ=0.625, P<0.001) (SUPPLEMENTARY TABLE 1). FDS reached borderline significance at this level of adjustment (ρ=0.319, P=0.035). In contrast, MDS-UPDRS Part IV item 4.1 (ρ=0.283, P=0.063) and item 4.2 (ρ=0.273, P=0.073) were attenuated and no longer significant. After additional adjustment for LEDD (Model B), the DK_50-PSQI association remained strong (ρ=0.577, P<0.001), while the FDS association was attenuated (ρ=0.257, P=0.096). Clinician-rated items remained non-significant in Models A and B (item 4.1, P=0.063-0.185; item 4.2, P=0.073-0.200). After adjustment for disease duration (Model C), the DK_50-PSQI partial correlation remained significant (ρ=0.580, P<0.001), whereas item 4.1 (ρ=0.202, P=0.200), item 4.2 (ρ=0.194, P=0.219), and FDS (ρ=0.251, P=0.108) were not significant.

In multivariable linear regression adjusting for age, sex, MDS-UPDRS Part III (Model A), and LEDD (Model B), DK_50 was the only independent predictor of PSQI (B=0.18, 95%CI 0.003-0.357, P=0.047). FDS (B=0.40, 95%CI -0.03 to 0.83, P=0.070), and MDS-UPDRS Part IV item 4.1 (P=0.187), and item 4.2 (P=0.275) did not reach significance (TABLE 2). After adjustment for disease duration (Model C), DK_50 attenuated to borderline non-significance (B=0.18, 95%CI -0.001 to 0.356, P=0.051), with FDS (P=0.063), item 4.1 (P=0.104) and item 4.2 (P=0.142) non-significant.

**TABLE 2.** Multivariable linear regression of PSQI on dyskinesia measures across covariate adjustment models.

| Model | Covariates | n | B | SE | $\beta$ | 95% CI | P |
| --- | --- | --- | --- | --- | --- | --- | --- |
| **MDS-UPDRS Part IV item 4.1** |  |  |  |  |  |  |  |
| Model A | Age + Sex + MDS-UPDRS Part III | 47 | 1.315 | 0.555 | 0.343 | 0.195 to 2.436 | 0.022 |
| Model B | Model A + LEDD | 47 | 0.834 | 0.621 | 0.218 | -0.421 to 2.089 | 0.187 |
| Model C | Model B + Disease duration | 47 | 1.076 | 0.647 | 0.281 | -0.231 to 2.384 | 0.104 |
| **MDS-UPDRS Part IV item 4.2** |  |  |  |  |  |  |  |
| Model A | Age + Sex + MDS-UPDRS Part III | 47 | 1.350 | 0.607 | 0.319 | 0.125 to 2.575 | 0.032 |
| Model B | Model A + LEDD | 47 | 0.769 | 0.696 | 0.182 | -0.636 to 2.174 | 0.275 |
| Model C | Model B + Disease duration | 47 | 1.109 | 0.741 | 0.262 | -0.388 to 2.606 | 0.142 |
| **PKG DK_50** |  |  |  |  |  |  |  |
| Model A | Age + Sex + MDS-UPDRS Part III | 47 | 0.211 | 0.091 | 0.336 | 0.028 to 0.394 | 0.025 |
| Model B | Model A + LEDD | 47 | 0.180 | 0.088 | 0.286 | 0.003 to 0.357 | 0.047 |
| Model C | Model B + Disease duration | 47 | 0.177 | 0.088 | 0.282 | -0.001 to 0.356 | 0.051 |
| **PKG FDS** |  |  |  |  |  |  |  |
| Model A | Age + Sex + MDS-UPDRS Part III | 47 | 0.522 | 0.211 | 0.380 | 0.096 to 0.949 | 0.018 |
| Model B | Model A + LEDD | 47 | 0.399 | 0.214 | 0.290 | -0.033 to 0.831 | 0.070 |
| Model C | Model B + Disease duration | 47 | 0.410 | 0.215 | 0.298 | -0.024 to 0.845 | 0.063 |

MDS-UPDRS, Movement Disorder Society-Unified Parkinson's Disease Rating Scale; PSQI, Pittsburgh Sleep Quality Index; PKG, Parkinson's KinetiGraph; DK\_50, median dyskinesia score; FDS, fluctuation dyskinesia score; LEDD, levodopa equivalent daily dose; B, unstandardised regression coefficient; SE, standard error; $\beta$ , standardised regression coefficient; CI, confidence interval.

## Discussion

Sleep disturbance and dyskinesia are two of the most disabling problems patients with PD describe^4,6^, yet their association has been characterised almost exclusively through brief, intermittent clinical observation. We show that the choice of dyskinesia measurement method substantially changes the apparent dyskinesia-sleep relationship in PD. To our knowledge, this is the first head-to-head comparison of continuous wearable and intermittent clinician-rated dyskinesia as correlates of subjective sleep quality, and the first in a demographically diverse UK cohort. The findings have implications for how persistent sleep complaints in PD are evaluated and how dyskinesia outcomes are specified in trials.

Mao et al.^12^, in 425 patients, found that subjective sleep quality and sleep latency were the PSQI components most strongly associated with dyskinesia. Tang et al.^13^ prospectively showed that high PSQI predicted incident LID at 36 months. Alongside with our findings, this evidence supports a bidirectional dyskinesia-sleep relationship where poor sleep impairs the cortical synaptic-downscaling process that protects against the maladaptive corticostriatal long-term potentiation underlying LID, while ongoing dyskinesia, particularly evening and nocturnal movement, directly fragments sleep. Our cross-sectional design cannot establish the direction, but both pathways are biologically plausible. One implication is that interventions targeting either domain alone may yield smaller benefits than combined approaches.

The methodological asymmetry between continuous and intermittent assessment likely explains the attenuation pattern. Clinic ratings sample a brief window of an inherently fluctuating phenomenon. The dyskinetic motor activity most relevant to sleep architecture occurs in the late afternoon, evening, and night, when clinic visits cannot capture it^17,29^. The strengthening of the DK_50-PSQI association after severity adjustment is consistent with negative confounding, whereby OFF-state-dominant patients with little dyskinesia obscured the dyskinesia-specific signal in unadjusted analysis.

Two further observations argue against this being a chance association. In patients followed before any LID emerged, the magnitude of the overnight decline in slow-wave activity predicted time to dyskinesia onset^27^, a pattern paralleled in the 6-OHDA rat model^26^. Once dyskinesia is established, raised morning cortical theta further characterises the dyskinetic state^28^. DK_50 is the median dyskinesia score across seven days, which makes it less a snapshot of any single dose cycle and more a reflection of the levodopa long-duration response, the slower, plasticity-driven motor effect that accumulates over days to weeks. That timescale aligns with what the PSQI assesses: sleep over the preceding four weeks. The fluctuation dyskinesia (FDS) captures shorter-timescale variability, which may be part of why it tracks PSQI less closely.

That the FDS-PSQI association weakened once LEDD was accounted for suggests the fluctuation component is partly carried by medication load rather than reflecting an independent neurobiological signal^29^. The attenuation by disease duration is harder to interpret cleanly, and three readings are compatible with our data. It may be a common-cause confounder, since both LID risk and sleep disturbance rise with disease progression^4,8^, so adjusting for it removes shared variance. Alternatively, it may sit closer to the mechanism itself, standing in for the cumulative corticostriatal remodelling that drives both phenomena in which case adjusting for it constitutes over-adjustment^31^. A third reading is more prosaic: with 47 patients and five covariates, we are at the edge of what the data can support, and the near-identical point estimates before and after adjustment are consistent with loss of power rather than abolition of the effect.

Persistent sleep complaints in PD are commonly attributed to nocturia, REM sleep behaviour disorder, restless legs syndrome, or depression^1,5^ but dyskinesia is rarely interrogated systematically. In a UK PD service introducing routine PKG monitoring, wearable-identified sleep disturbance featured at 58% of follow-up appointments and dyskinesia was detected in patients who had not previously reported it^30^. Our data support incorporating wearable dyskinesia assessment into the care of patients with PD and persistent poor sleep, particularly when clinic-based dyskinesia screening is negative. For research, the divergent behaviour of DK_50 versus FDS suggests that interventional trials testing whether dyskinesia optimisation improves sleep should use time-averaged rather than fluctuation-based dyskinesia outcomes, combined with objective sleep measurement to allow inferences about direction. Given the bidirectional dyskinesia-sleep relationship suggested by the combined cross-sectional and prospective evidence, future trials should test whether combined interventions (dyskinesia optimisation plus sleep-targeted therapy) outperform either alone.

Several limitations apply. The cross-sectional design precludes causal inference, and the modest sample restricted covariate modelling, contributing to the borderline result after disease-duration adjustment. Disease duration was measured from formal diagnosis rather than symptom onset, which likely underestimates the true biological duration. Sleep was assessed only by self-report. The global PSQI obscures component-level patterns and cannot capture sleep architecture. Future studies should adjust for several variables that may influence both dyskinesia and sleep in PD. Depression and anxiety co-cluster with both and were not measured here. Cognition moderates self-reported sleep quality. Nocturia and restless legs syndrome fragment sleep independently of dyskinesia. The remaining MDS-UPDRS Part IV items (4.3-4.6) capture OFF-state dystonia and motor fluctuations that often co-occur with dyskinesia. Self-reported ethnicity also matters, given evidence of greater non-motor burden in non-White populations^32^. Strengths include continuous wearable dyskinesia measurement, pre-specified hierarchical adjustment, a demographically diverse cohort, and consistent findings across correlation and regression approaches.

## Conclusions

In PD, greater dyskinesia burden was associated with worse subjective sleep quality, but the strength of this association depended on how dyskinesia was measured. Wearable-derived DK_50 tracked PSQI across adjustment for age, sex, motor severity, and LEDD. Clinician-rated MDS-UPDRS Part IV items did not. The role of disease duration remains uncertain in this sample. The findings support assessing dyskinesia with continuous wearable monitoring in PD patients with persistent poor sleep, and pairing time-averaged dyskinesia outcomes with objective sleep measurement in future trials.

## Data Availability

De-identified participant data and supporting documents including study protocol, statistical analysis plan, data dictionary, and consent form will be made available after publication upon reasonable request to the corresponding author, subject to a data use agreement and applicable ethical constraints.

## Acknowledgments

We thank all participants for taking part in this study. We also thank the study project team and supporting staff for their assistance. This work was supported by a UK Research and Innovation Knowledge Transfer Partnership (KTP) award (Innovate UK, 2021-2022, Round 4).

## Authors’ roles

Research project:

A. Conception: VA, LPC, AJN, CS

B. Organization: VA, LPC AJN, CS

C. Execution: VA

Statistical Analysis:

A. Design: VA, JB

B. Execution: VA, JB

C. Review and critique: VA, EB, EC, KCD, AZ, KR, CQ, TB, CB, DG, JB, LPC, AJN, CS

Manuscript:

A. Writing of the first draft: VA

B. Review and critique: VA, EB, EC, KCD, AZ, KR, CQ, TB, CB, DG, JB, LPC, AJN, CS

## Funding Sources

This study was supported by UK Research and Innovation through a Knowledge Transfer Partnership (Innovate UK, 2021-2022, Round 4).

## Role of the Funder/Sponsors

The funders/sponsors had no role in study design, data collection, management, analysis, interpretation, manuscript preparation, review, approval, or the decision to submit for publication.

## Conflicts of Interest

All authors report no personal fees, equity holdings, or advisory compensation related to this work. AJN does not have conflict of interest or disclosures to report in relation to this manuscript but he reports in general, not related to this work, research grant support from Parkinson’s UK, Barts Charity, Cure Parkinson’s, the National Institute for Health and Care Research, Innovate UK, UKRI, HORIZON 2020, the Medical College of Saint Bartholomew’s Hospital Trust, Alchemab, Aligning Science Across Parkinson’s/Global Parkinson’s Genetics Program (ASAP-GP2), and the Michael J. Fox Foundation within the past three years. AJN has also received consultancy or personal fees from AbbVie, Bial, and Roche. LPC does not have conflict of interest or disclosures to report in relation to this manuscript but reports, unrelated to this work, research grant support from the Michael J. Fox Foundation, director and speaker fees for the RBD Symposium at Guy’s and St Thomas’ NHS Foundation Trust, consultancy fees from Takeda and Idorsia within the past three years.

## Ethical Compliance Statement

Ethical approval for this study was granted by the London-Dulwich Research Ethics Committee (reference: 23/PR/1526). All procedures were conducted in accordance with the principles of the Declaration of Helsinki. Written informed consent was obtained from all participants prior to enrolment. We confirm that we have read the position on issues involved in ethical publication and affirm that this work is consistent with those guidelines.

## Notes

### Competing Interest Statement

The authors have declared no competing interest.

### Author Declarations

The London-Dulwich Research Ethics Committee gave ethical approval for this work (reference 23/PR/1526)

