## Supplementary Table 1 for "Wearable Evidence Linking Dyskinesia Burden to Sleep Quality in Parkinson’s Disease"

**SUPPLEMENTARY TABLE 1. Associations between sleep quality and dyskinesia measures across covariate adjustment models**

| **Model** | **Covariate set** | **n** | **Partial ρ** | **P-value** |
| --- | --- | --- | --- | --- |
| **MDS-UPDRS Part IV item 4.1** | | | | |
| Unadjusted | (no covariates) | 47 | 0.386 | 0.007 |
| Model A | Age + Sex + MDS-UPDRS Part III | 47 | 0.283 | 0.063 |
| Model B | Model A + LEDD | 47 | 0.206 | 0.185 |
| Model C | Model B + Disease duration | 47 | 0.202 | 0.200 |
| **MDS-UPDRS Part IV item 4.2** | | | | |
| Unadjusted | (no covariates) | 47 | 0.392 | 0.006 |
| Model A | Age + Sex + MDS-UPDRS Part III | 47 | 0.273 | 0.073 |
| Model B | Model A + LEDD | 47 | 0.199 | 0.200 |
| Model C | Model B + Disease duration | 47 | 0.194 | 0.219 |
| **PKG DK_50** | | | | |
| Unadjusted | (no covariates) | 47 | 0.487 | <0.001 |
| Model A | Age + Sex+ MDS-UPDRS Part III | 47 | 0.625 | <0.001 |
| Model B | Model A + LEDD | 47 | 0.577 | <0.001 |
| Model C | Model B + Disease duration | 47 | 0.580 | <0.001 |
| **PKG FDS** | | | | |
| Unadjusted | (no covariates) | 47 | 0.265 | 0.072 |
| Model A | Age + Sex + MDS-UPDRS Part III | 47 | 0.319 | 0.035 |
| Model B | Model A + LEDD | 47 | 0.257 | 0.096 |
| Model C | Model B + Disease duration | 47 | 0.251 | 0.108 |

MDS-UPDRS, Movement Disorder Society-Unified Parkinson's Disease Rating Scale; PSQI, Pittsburgh Sleep Quality Index; PKG, Parkinson's KinetiGraph; DK_50, median dyskinesia score; FDS, fluctuation dyskinesia score; LEDD, levodopa equivalent daily dose; ρ, Spearman's rank correlation coefficient.
